# SARS-CoV-2 infections among personnel providing home care services for the elderly in Stockholm, Sweden

**DOI:** 10.1101/2020.12.18.20248511

**Authors:** Sadaf Sakina Hassan, Åsa Seigerud, Laila Sara Arroyo Mühr, Sara Nordqvist Kleppe, Elisa Pin, Anna Månberg, Sophia Hober, Peter Nilsson, Lars Engstrand, K. Miriam Elfström, Jonas Blomqvist, Kalle Conneryd Lundgren, Joakim Dillner

## Abstract

**Background:** In March 2020, Stockholm, Sweden was hit by a severe outbreak of SARS-CoV-2. Four weeks later, a systematic study of testing for past or present infections among healthcare workers in the region was launched. Only a minority of COVID-19 related deaths occurred at hospitals and the study was therefore extended to employees in companies providing home care services for the elderly.

**Methods:** Five companies offered participation to 438 employees at work and 405 employees (92.5%) were enrolled. Serum samples were analyzed for IgG to SARS-CoV-2 and throat swabs were tested by for the SARS-CoV-2 virus by PCR.

**Results:** Among home care employees, 20.1% (81/403) were seropositive, about twice as many as in a simultaneously enrolled reference population (healthcare workers entirely without patient contact, n=3,671; 9.7% seropositivity). Only 13/379 employees (3.4%) had evidence of a current infection (PCR positivity). Among these, 5 were also seropositive (a sign of past infection or lingering infection after symptoms have resolved) and 3 were positive with only low amounts of virus. The combination of high amounts of virus and no antibodies, a characteristic for pre-symptomatic COVID-19, was thus present only in 5 employees (1.3%).

**Conclusions:** Personnel providing home service for the elderly appear to be a risk group for SARS-CoV-2 infection. Employees likely to be pre-symptomatic for COVID-19 can be readily identified by screening. Increased attention for protection of employees as well as of the elderly they serve is warranted.

## Introduction

The severe acute respiratory syndrome coronavirus 2 (SARS-CoV-2) has since its first report in Wuhan, China in December 2019 escalated to a pandemic, affecting all countries around the world [1, 2]. As of 23^rd^ November 2020, over 61.8 million confirmed Coronavirus Disease 2019 (COVID-19) cases caused by SARS-CoV-2, and over 1.4 million deaths have been reported worldwide [3]. The older segment of the population (>70 years of age) is a particularly vulnerable group. They have been shown to be more susceptible to severe infections and thus are at higher risk of dying due to SARS-CoV-2 infection [4, 5].

Several public health measures have been introduced to prevent and control the spread of the virus in different countries [6]. Countries such as France, Spain, and Italy declared total lock downs in March 2020, while other countries like South Korea and Sweden had different approaches to control the spread without opting for lock downs [7]. However, all countries without exception have implemented strategies to protect the elderly from SARS-CoV-2 infections.

The Public Health Agency of Sweden has at the time of writing, reported more than 200,000 confirmed SARS-CoV-2 cases and more than 7,000 deaths due to COVID-19 in Sweden. People aged over 70 years old comprise about 25,000 confirmed cases (12% of total confirmed cases) and almost 90% of total deaths due to COVID-19 in Sweden (Folkhälsomyndigheten.se data, accessed on Nov 23rd, 2020). The Swedish government introduced a protection order for residential care centers and the Public Health agency of Sweden advised against visits to the elderly, neither at home nor at residential care centers [8]. Many reports have indicated a higher prevalence of SARS-CoV-2 virus among health care personnel compared to the general public [9]. A study from Stockholm, Sweden, screened elderly care personnel with rapid detection of IgG against SARS-CoV-2 and showed that 20.3% of employees were tested positive [10]. In Sweden, while some persons over age 70 live in residential care centers, most of them receive home care services instead, meaning that services and personal care are provided regularly at home. In January 2020, almost 80,000 persons were living at residential care centers, while about 190,000 people over age 70 had received home care services in Sweden [11]. Hence, it is of great value to gain knowledge on the spread of SARS-CoV-2 among home care services personnel in order to design strategies to control the infection among this group and the personnel working with them.

The aim of the study was to investigate past (serum antibody positivity) or current (presence of viral RNA) SARS-CoV-2 infection in personnel working for elderly home care services during the COVID-19 outbreak.

## Material and Methods

### Subjects and samples

Employees that were on duty from five different elderly home care service companies (n=438) in the region of Stockholm were invited to participate in the study during the period 11^th^ May-17^th^ June 2020. The number of employees at the companies ranged from 27-291 (Table 1).

**Table 1:** Characteristics of health care workers at elderly home care services and samples collected.

| **Table 1: Characteristics of employees at elderly home care services and samples collected** |  |  |  |  |  |  |  |
| --- | --- | --- | --- | --- | --- | --- | --- |
| **Home care service company** | **Number of**<br>**total employees** | **Number of employees**<br>**participating in the study** | **Number of**<br>**swabs collected** | **Number of**<br>**blood samples** | **Number of**<br>**men** | **Number of**<br>**women** | **Median Age**<br>**Total**<br>**(men/women)** |
| **Hemtjänst 24 Omsorg** | 28 | 28 | 28 | 28 | 14 | 14 | 32 (32.5/32) |
| **Hägersten-Liljeholmen-Älvsjö Hemtjänst** | 291 | 271 | 261 | 270 | 126 | 145 | 43 (44/43) |
| **Roo Hemtjänst AB** | 72 | 67 | 65 | 67 | 35 | 32 | 43 (42/44) |
| **Saras Omsorg** | 20 | 18 | 16 | 18 | 10 | 8 | 37 (42.5/35) |
| **Vardaga Hemtjänst** | 27 | 21 | 17 | 20 | 7 | 14 | 44 (44/40) |
| **All companies** | 438 | 405 | 387 | 403 | 192 | 213 | 43 (42.5/40) |

The participants were asked to provide a throat swab sample for SARS-CoV-2 RNA detection and a blood sample to perform serological analysis of antibodies for SARS-CoV-2.

Written informed consent was obtained from all participants in the study. The study was approved by the National Ethical Review Agency of Sweden (Decision number 2020-01620 and 2020-01881). Trial registration number: ClinicalTrials.gov NCT04411576. All methods were carried out in compliance with the Helsinki declaration.

As described in a parallel paper [12], we assembled a reference group of 3671 hospital employees that had no patient contact whatsoever. There were simultaneously enrolled at the Karolinska University hospital, also in Stockholm, Sweden. The Hospital employees without patient contact were, for example, working with administration, economy, information technology, engineering, research & innovation, or at the hospital laboratories. As hospital employees are very motivated to participate (92% participated in the parallel study) [12] we considered that use of hospital employees without patient contact would be a rapid strategy to obtain a large reference group with minimum selection bias because of non-participation that could be considered as approximately representative of the underlying degree of SARS-CoV-2 exposure in the region.

### SARS-CoV-2 nucleic acid detection

The participants were requested to give a throat swab sample as described in the user manual [13]. The samples were inactivated by incubation at 75 degrees for 50 minutes. The viral RNA was extracted according to the manufacturers protocol by performing MGISP-960 automated extraction standard workflow with MGIEasy Magnetic Beads Virus DNA/RNA extraction kit (Wuhan MGI Tech Co, Ltd). Real time polymerase chain reaction was performed on QuantStudio5 instruments and software (Design and Analysis Software v1.5.1, Thermo Scientific), using the BGI 2019-nCoV detection kit (BGI Real-Time RT-PCR for detecting 2019 nCoV) according to the manufacturers protocol along with internal parameters for testing process and sampling quality. Every step of the laboratory work followed validated standard operating protocols for reproducibility, sensitivity, specificity, and lack of cross-reactivity with other strains of coronavirus. For the sample preparations, the safety routines according to BSL2 requirements were followed. Results of the PCR tests were classified as positive, negative, or inconclusive/censored.

### Serological analysis of antibodies for SARS-CoV-2

Blood was collected in tubes containing serum separating gel and centrifuged at 2000 × g for 10 minutes at room temperature to collect the serum. To inactivate the samples, they were incubated at 56 degrees for 30 minutes before being stored at minus 20 degrees until further analysis. Sera were analyzed against three different variants of viral proteins which included: Spike trimers that contain prefusion-stabilized spike glycoprotein ectodomain [14], Spike S1 domain and Nucleocapsid protein.

The serum samples were analyzed with FlexMap3D instruments (Luminex Corp) using a multiplex antigen bead array in a high-throughput 384-plate format [15]. Samples were assigned IgG positive if the sera were reactive against at least two of the three viral antigens. For each antigen, the cut-off for seropositivity was defined as mean +6SD of 12 negative control samples that were included in each analysis run.

The serology assay was evaluated based on the analyses of 243 samples from COVID-19 convalescents (defined as PCR-positive individuals sampled more than 16 days after positive PCR test) and 442 negative samples (defined as samples collected 2019 or earlier in the same region, including 26 individuals with confirmed infections of other Coronaviruses than SARS-CoV-2). The assay had a 99.2% sensitivity and 99.8% specificity based on these samples.

Results are presented as descriptive statistics examining the prevalence of antibodies and viral infection by home care service company.

## Results

A total of 438 employees in five elderly home care service companies were invited and 405 employees (92.5%) were enrolled in the study. The participants provided 387 swab throat samples and 403 serum samples between 11^th^ May-17^th^ June 2020. In total, 375 participants provided both swab and blood samples with valid results (Fig 1). The mean age of the participants was 43 years. The median age ranged between 32 to 44 at the different home care companies (Table 1).

**Figure 1:**
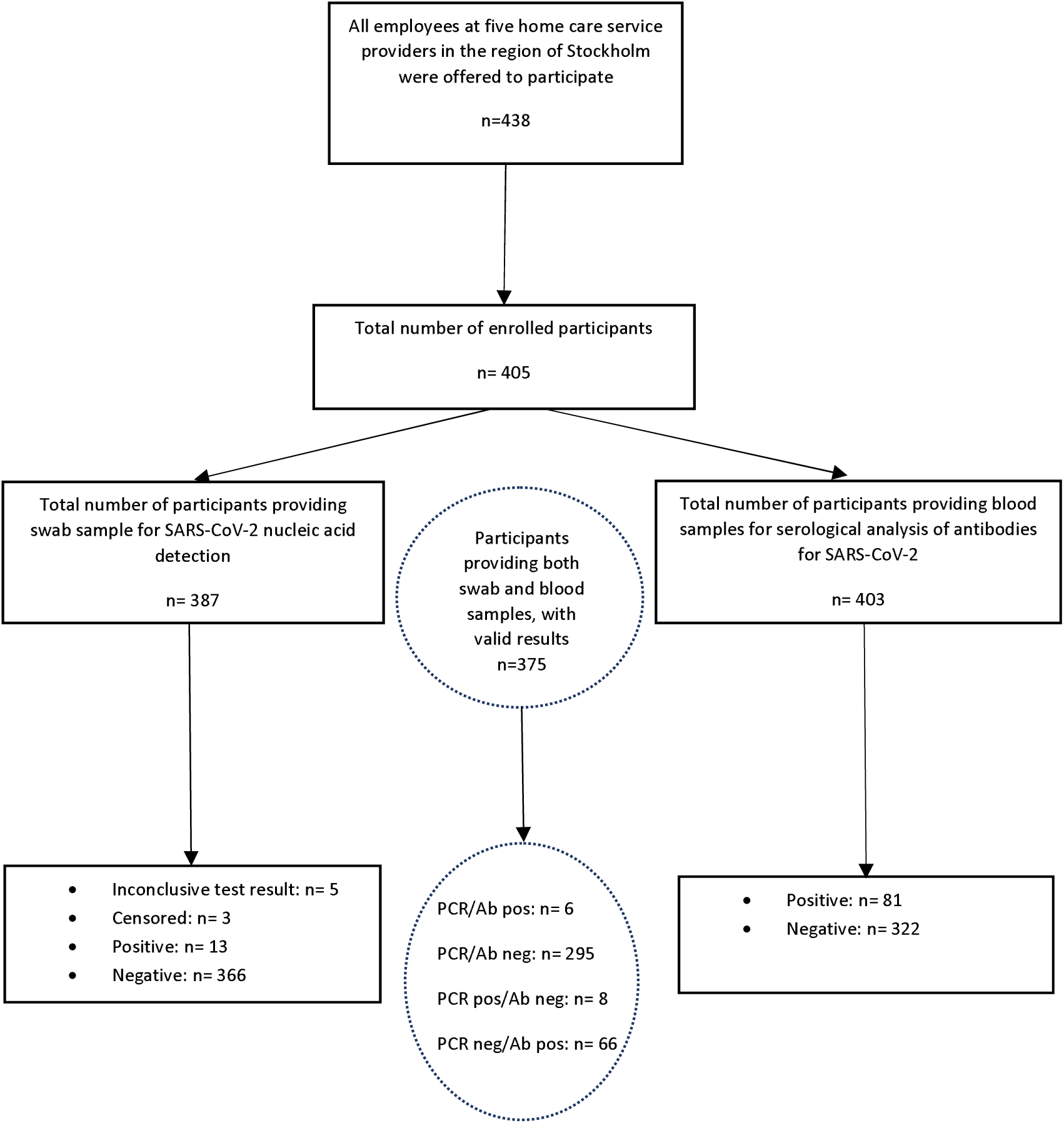
Flow Chart summarizing the screening of elderly care personnel for nucleic acid detection and serological analysis of antibodies SARS-CoV-2 Results of the PCR test were classified as positive, negative, or inconclusive/censored. Results were censored if a repeat sample had been taken and the results differed. Inconclusive means that the test result was ambiguous. PCR=Polymerase Chain Reaction, Ab=antibodies

### SARS-CoV-2 nucleic acid detection in elderly care personnel

The overall prevalence of confirmed SARS-CoV-2 by real time PCR was 3.4% (13/379 participants), with 2 out of 5 companies having no PCR confirmed case of SARS-CoV-2 at all (Table 2). The prevalence of confirmed PCR positivity was higher among men (6.1%, n=11) compared to women (n=2) (Table 2).

**Table 2:**
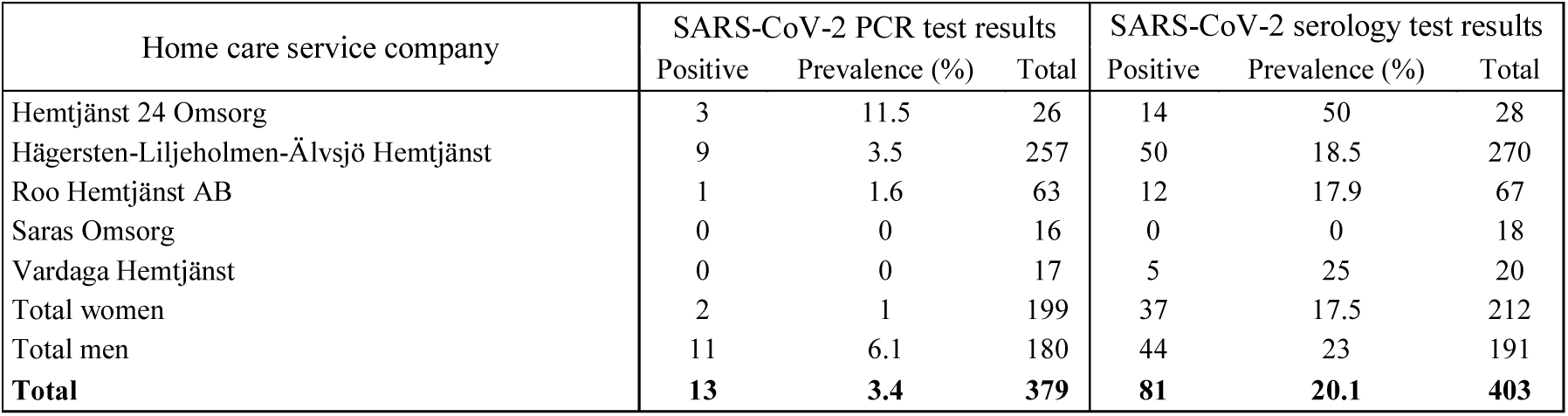
Results of SARS-CoV-2 PCR and serology testing from employees at elderly home care service companies.

| **Table 2: Results of SARS-CoV-2 PCR and serology testing from employees at elderly home care services** |  |  |  |  |  |  |
| --- | --- | --- | --- | --- | --- | --- |
| Home care service company | SARS-CoV-2 PCR test results |  |  | SARS-CoV-2 serology test results |  |  |
|  | Positive | Prevalence (%) | Total | Positive | Prevalence (%) | Total |
| Hemtjänst 24 Omsorg | 3 | 11.5 | 26 | 14 | 50 | 28 |
| Hägersten-Liljeholmen-Älvsjö Hemtjänst | 9 | 3.5 | 257 | 50 | 18.5 | 270 |
| Roo Hemtjänst AB | 1 | 1.6 | 63 | 12 | 17.9 | 67 |
| Saras Omsorg | 0 | 0 | 16 | 0 | 0 | 18 |
| Vardaga Hemtjänst | 0 | 0 | 17 | 5 | 25 | 20 |
| Total women | 2 | 1 | 199 | 37 | 17.5 | 212 |
| Total men | 11 | 6.1 | 180 | 44 | 23 | 191 |
| **Total** | **13** | **3.4** | **379** | **81** | **20.1** | **403** |

### Serological analysis of antibodies for SARS-CoV-2

The total seroprevalence among the elderly care personnel was 20.1% (81/403 employees were seropositive). One elderly home care service company had 14/28 employees showing antibody positivity to SARS-CoV-2, but another home service company did not have any seropositive employees at all (Table 2).

By comparison, the simultaneously enrolled reference group of employees without patient contact (n=3671) had a 9.7% seropositivity rate when tested by exactly the same serum test (OR for seropositivity when employed in home care services compared to the reference group: 2.3 (95% CI 1.8-3.1), p<0.0001, [12]).

Among the 13 SARS-CoV-2 PCR positive home services employees, eight were negative for SARS-CoV-2 antibodies. Five of these participants had Cycle Threshold values (Ct) lower than 27, indicating a strong positivity for SARS-CoV-2 (data not shown).

## Discussion

We find that personnel working in companies providing home services for the elderly are a risk group for SARS-CoV-2 infection, with an about twice as high risk for seropositivity as the reference group. The comparatively high seroprevalence in combination with a rather low proportion of PCR-positive subjects in our study suggests that most of the first wave of SARS-CoV-2 infections had already started to subside when the study was performed. This is well in line with the statistics showing a sharp peak of COVID-19 -related death in the region in early April [16]. Indeed, a substantial proportion of PCR-positive subjects either were seropositive or had very low amounts of virus (signs of a previous infection or a lingering infection after resolution of symptoms).

Only 5/405 individuals had the typical pre-symptomatic pattern of being PCR positive with high amounts of virus in combination with lack of antibodies. Pre-symptomatic subjects are potential “super spreaders” [17]. We thus find that in a rather large cohort of home service employees there existed only a small number of potentially super-spreading individuals. These were easily identified by the screening and the fact that they were identified among subjects who serve multiple elderly clients suggests that such identification of potential super-spreaders could be considered.

A strength of our study was that it was performed early on, at a time when large-scale testing was not available in Sweden and that we exploited high performance assays in experienced academic laboratories. Although we find that the first wave of the epidemic was already declining in the region there was still evidence of actively ongoing transmission at that time. Furthermore, a large-scale reference group without any contact with patients could be assembled at the same time and used for comparison.

Weaknesses of the study include the fact that only 5 companies providing home care services participated in the study. There are 12 times as many companies providing home care services in the region and the companies that participated were not selected at random, but participation was decided on a first-come, first-serve basis (only a minority of the companies had decided to participate when the start date of the study was imminent). Also, the elderly who received the home care services were not enrolled, limiting our ability to document whether services provided by infectious home services personnel may have transmitted infections to the users. We did not enroll the elderly because of issues regarding complexity in obtaining a fully informed and coercion-free consent and issues regarding requirements for documentation of testing results of the users, issues that were not readily resolved.

Our findings on the seropositivity among home care service employees are similar to another study [10], implying that there are now independent studies pointing to that this is a risk group for SARS-CoV-2 infection.

## Conclusion

In summary, we find that employees in home care services are a risk group for contracting SARS-CoV-2 infection, as evidenced by a higher seroprevalence compared to the reference group. We also find that there were only a few employees with the typical testing result of pre-symptomatic subjects, who may be potential “super spreaders”.

Our findings suggest that the personnel providing home care services appear to be a risk group of SARS-CoV-2 infection. Increased attention for protection and screening of personnel as well as of the elderly they serve is warranted.

## Data Availability

Non applicable

## Conflict of interest

All authors declare no conflict of interest.

## Acknowledgements

Authors would like to thank Camilla Lagheden, Carina Eklund, Suyesh Amatya, Eni Andersson, Helena Andersson, Sofia Bergström, Emine Eken, Pedram Farsi, Cecilia Hellström, Yasmin Hussein, Roxana Merino Martinez, Jennie Olofsson, Björn Pfeifer, Ulla Rudsander, Balazs Szakos, Hanna Tegel, and Emel Yilmaz for excellent technical assistance. This research was supported by the County Council of Stockholm.

## Notes

### Competing Interest Statement

The authors have declared no competing interest.

### Clinical Trial

NCT04411576

### Author Declarations

The study was approved by the National Ethical Review Agency of Sweden (Decision number 2020-01620 and 2020-01881). Trial registration number: ClinicalTrials.gov NCT04411576. All methods were carried out in compliance with the Helsinki declaration. Written informed consent was obtained from all participants in the study.

